# NO INCREASE IN RELATIVE MORTALITY RATES FOR THOSE WITHOUT A COLLEGE DEGREE DURING COVID-19: AN ANOMALY

**DOI:** 10.1101/2021.07.20.21260875

**Authors:** Anne Case, Angus Deaton

## Abstract

American mortality rates have diverged in recent years between those with and without a four-year college degree, and there are many reasons to expect the education-mortality gradient to have steepened during the pandemic. Those without a BA are more likely to work in frontline occupations, to rely on public transportation, and to live in crowded quarters, all of which are associated with an increase in infection risk, a risk that was zero prior to the pandemic. We use publicly available data from the National Center for Health Statistics on deaths by age, sex, education and race/ethnicity to assess the protective effect of a BA in 2020 compared to 2019. While the BA was strongly protective during 2020, the ratio of mortality rates between those with and without a degree was little changed relative to pre-pandemic years. Among 60 groups (gender by race/ethnicity by age) that are available in the data, the relative risk reduction associated with a BA *fell* for more than half the groups between 2019 and 2020, and increased by more than 5 percentage points for only five groups. Our main finding is not that the BA was protective against death in 2020, which has long been the case, but that the protective effect was little different than in 2019 and earlier years, in spite of the change in the pattern of risk by occupation and income. The virus maintained the mortality-education gradient that existed pre-pandemic, at least through the end of 2020. Our results suggest that changes in the risk of infection were less important in structuring mortality than changes in the risk of death conditional on infection.

## 1. Introduction

The bachelor’s (BA) degree has increasingly divided the mortality experience of Americans, especially since 2010. Life expectancy at age 25 increased by 1.5 years from 2010 to 2018 for those with a BA, while decreasing by 1 year for those without; by 2018, the gap had risen to a historical high of six years, Case and Deaton (2021). The COVID-19 pandemic was far from an equal opportunity killer, and there were sharply different mortality rates by race/ethnicity, CDC (2021), Andrasfay and Goldman (2021a, b) Woolf et al (2021) and by education, documented in California between March and October 2020, Chen YH et al (2020) and with interactions between education and race/ethnicity for the US as a whole in 2020, Chen JT et al (2021).

In this paper we focus on the protection provided by a BA degree during 2020 compared with 2019 and earlier years. We measure the protection from the BA analogously to the efficacy of a drug or vaccine, as the difference in deaths between those without and with the degree, divided by the deaths among those without the degree. This is a mechanical calculation, not an attribution of causality; those with and without a degree are different in many ways including their pre-existing risk factors, co-morbidities, occupations, transportation and living arrangements, all of which affect their probability of being infected or of dying once infected, and it is these differences, not the degree itself, that structure risk. In line with this, we use the term efficacy as a shorthand for the percentage reduction in mortality associated with the BA degree.

A fatal infectious disease could, in principle, narrow educational differences in mortality if everyone were to have an equal chance of being infected and of dying. But infection risks for COVID-19 differ by occupation; many highly educated people were able to work safely from home, while those who worked as frontline workers, in retail, services, transportation, warehousing, or meatpacking ran the risk of infection. Blau et al (2020) find that “frontline workers are disproportionately comprised of less educated and disadvantaged minority workers, especially Hispanics, and immigrants, and earn below average wages, with a substantial share of workers in the bottom wage quartile. These workers, even healthcare workers, now face much higher risks than traditionally incurred in these occupations.” Daly et al (2020) note “education is a common thread contributing to large differences in the severity of the virus’s impact across individuals.” That excess mortality was indeed higher in frontline occupations is documented for California by Chen YH et al (2021). Less-educated Americans are also more likely to have low incomes and to live in more crowded or inferior housing and are less likely to be able to avoid contact with others.

Our hypothesis is that the more educated would not only be less likely to die, as has long been the case, but that the educational gradient in relative mortality rates should have increased during the pandemic.

We find that while the BA was strongly protective during 2020, the ratio of mortality rates between those with and without a degree was little changed relative to pre-pandemic years. Among the 60 groups (gender by race/ethnicity by age) that are available in the data, the efficacy of a BA *fell* for more than half the groups between 2019 and 2020, and increased by more than 5 percentage points for only five groups. Our main finding is not that the BA was protective against death in 2020, but that the protective effect was little different than in 2019 and earlier, in spite of the change in the pattern of risk by occupation and income.

## 2. Data and methods

We use publicly-available data provided by the Centers for Disease Control and Prevention (2021) on deaths from COVID-19 in 2020 and deaths from all causes in 2019 and 2020 aggregated by race/ethnicity, by broad age-groups, by sex, by educational level and by year. We combine the two categories below BA (High School or less and some college) into a non-BA category, and attribute education to deaths with unknown education (2%) so as to maintain the fractions with a BA in each age, sex, and race/ethnicity category. We work with six ethnic/racial groups, Hispanics, non-Hispanic Whites (NHW), non-Hispanic Blacks (NHB), non-Hispanic Asians (NHAsian, including non-Hispanic Hawaiian Natives and other Pacific Islanders), non-Hispanic American Indians and Alaskan Natives (NHAIAN) and those who report two or more races (NHMany); we drop the unknown (other) category. We exclude those aged 24 or younger—for whom the BA distinction is not meaningful—leaving age groups 25−39, 40−54, 55−64, 65−74, and 75 plus. There are 60 (6×2×5) groups in all. For most presentations, we work with two age groups, 25−64, and 65 plus.

Population counts by BA/non-BA are taken from the American Community Survey via IPUMS USA database, Ruggles et al (2021); the 2019 numbers are used unchanged for 2020. For earlier years, 2010-2018, we use NVSS Mortality Multiple Cause of Death Files from the Vital Statistics Online Data Portal (2021).

We analyze the difference between 2020 and 2019 mortality rates by sex, age, education, and race/ethnicity. This difference is of interest in and of itself, and corresponds to calculations of the change in life expectancy from 2019-2020 by Andrasfay and Goldman (2021a, b) and by Woolf et al (2021). Many earlier studies have analyzed *excess* mortality during the epidemic, comparing actual deaths with those expected, the latter estimated from models using data from earlier years or months. There is no uncontroversial way of forecasting mortality; for example, accidental drug overdoses were rising before the pandemic, and rose more rapidly during it, which may or may not have been caused by the pandemic. Moreover, we would need forecasts for all population groups, including small but important groups, such as AIAN; examination of the data show that such forecasts are difficult with small population sizes. Comparisons of mortality in 2020 and 2019 are straightforward and transparent. The Centers for Disease Control and Prevention (CDC) estimate that over the period from late January 2020 to late February 2021, approximately 75%-88% of excess deaths were directly related to COVID-19, Rossen et al (2021). If we were to interpret the increase from 2019 to 2020 as a measure of excess mortality, we would have a ratio of excess to reported COVID-19 mortality of 1.28 from March to December 2020, consistent with the CDC range and with previous estimates for the year, Woolf et al (2021).

## 3. Results

Figure 1 shows the percent of each sub-population with a BA degree in 2019, by race/ethnicity, by gender, and by age, for two broad age groups (25-64, and 65 and above). Black bars denote men; gray bars, women. Overall, a third of adult Americans have a four-year degree, but there are large differences by race/ethnicity and age. Educational attainment has increased over time, so that, except for NHAIAN men, rates are higher for the young. Rates are highest (60 percent) for young NHAsians, followed by NHW and NHMany and are lowest among NHB, Hispanics, and NHAIAN.

**Figure 1.**
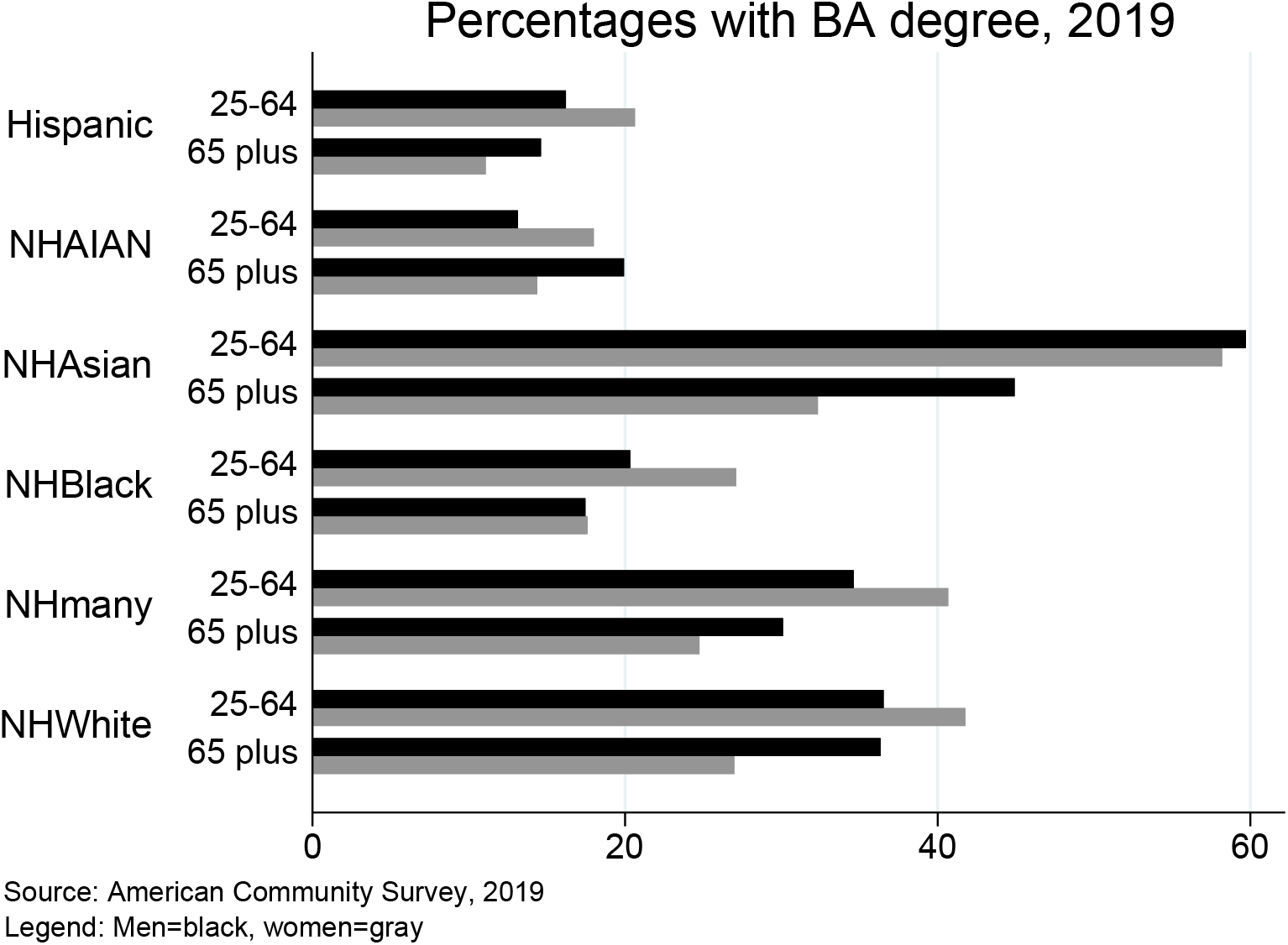

Figure 2 shows the percentage by which mortality, *m*, is lower among those with the BA degree, over the period from 2005 to 2019, for all groups together and for five of the six ethnic groups, over two broad age groups, for men and women taken together; we do not adjust for age within the two age categories. Each line in Figure 2 shows, for each group in each year, the efficacy of a BA, defined as 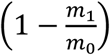 where *m*_1_ and *m*_0_ are the mortality rates for those with and without the degree.

**Figure 2.**
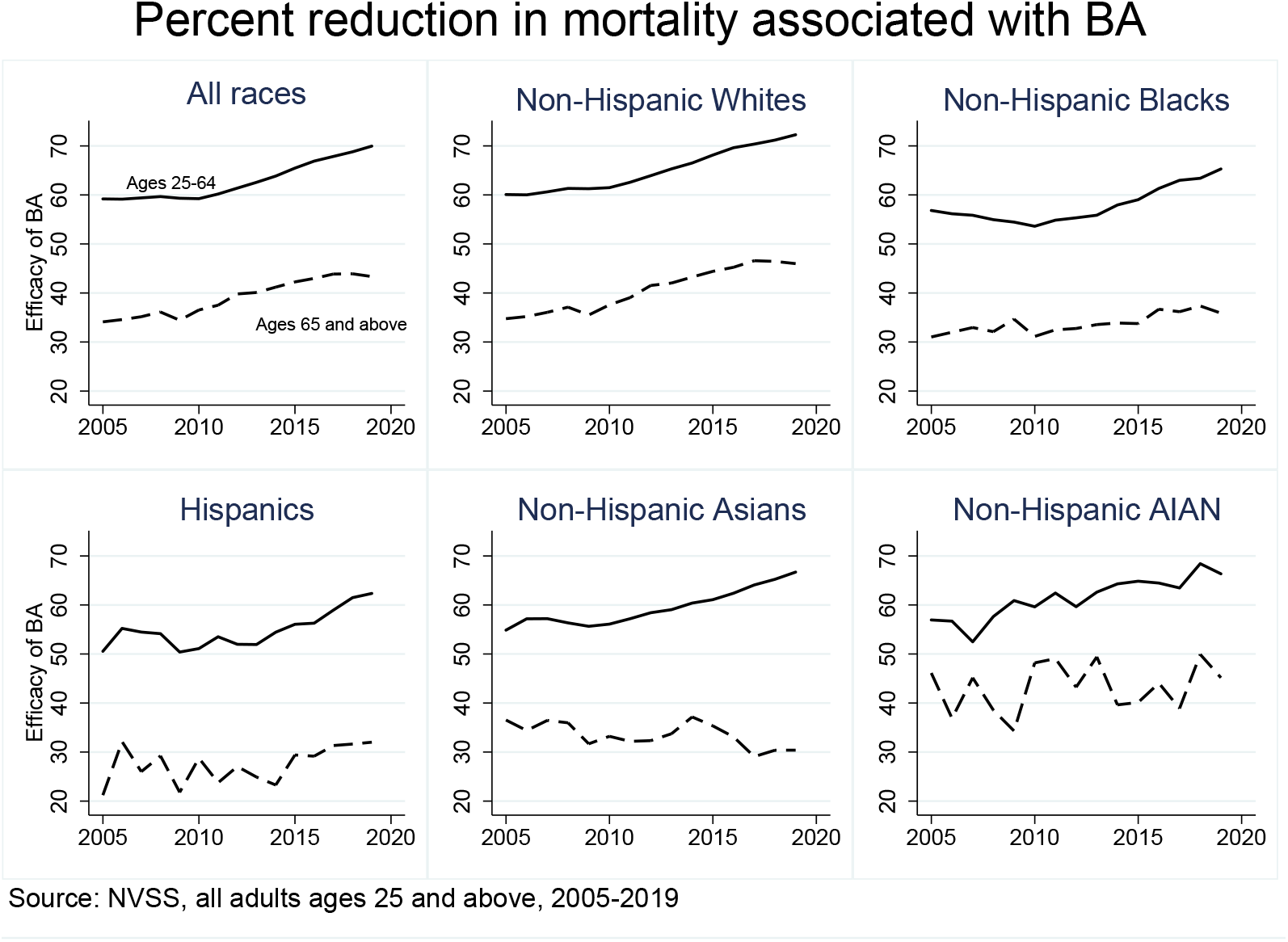

For ages 25−64, the efficacy of the BA has risen, particularly since 2010. The smallest effect is for Hispanics, but efficacy for all groups was more than 60 percent in 2019. Efficacy is weaker for the elderly, but has also grown over time for NHW, NHB and Hispanics; by 2019, the mortality advantage associated with a BA was between 30 and 50 percent for all older groups. (The bottom right panel reflects the small numbers of NHAIANs. A version of Figure 2 with five age groups shows similar patterns, with efficacy falling with age.

Figure 3 shows patterns of mortality rate increase between 2019 and 2020. We separate by broad age groups, gender, BA status, and the six races/ethnicities. We label each pair of bars with the percentage by which the BA increase in the mortality rate is lower than the non-BA increase, e.g., for younger NHAIAN the increase in the mortality rate from 2019 to 2020 was 68% lower for those with a BA. These numbers are comparable to the efficacy numbers in Figure 2 and, once again, they are always lower for the older group. The Figure replicates the now-standard finding that Hispanic, NHB and, especially, NHAIAN Americans have suffered disproportionately during 2020, with NHW, NHMany, and NHAsian Americans affected the least, though every group experienced increased mortality.

**Figure 3.**
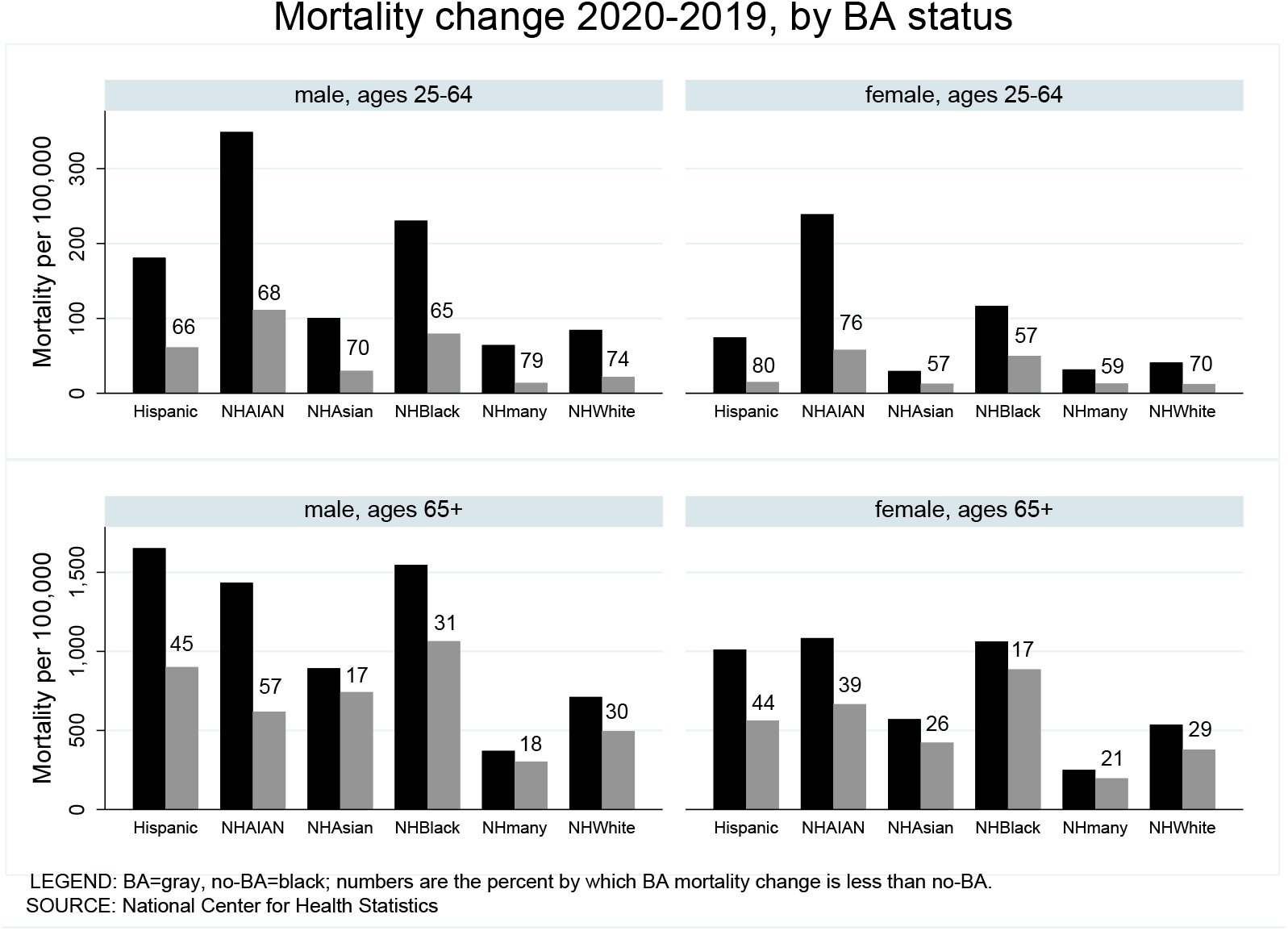

Figure 4 moves to increases in *relative* risk, where the patterns are quite different. For each of the 48 groups shown, we show the *ratio* of the increase in the mortality rate from 2019 to 2020 to their mortality rate in 2019. For the whole population, the ratio was 0.17; within the groups, it ranges from a high of 0.52 for younger Hispanic men without a BA to a low of 0.09 for younger adult NHW women irrespective of BA status. The racial/ethnic patterns single out Hispanics as having seen the largest increase in relative risk, not NHAIAN, whose large increase in mortality is now compared to their long-standing high mortality rate. The low relative risk of the WNH community is notable.

**Figure 4.**
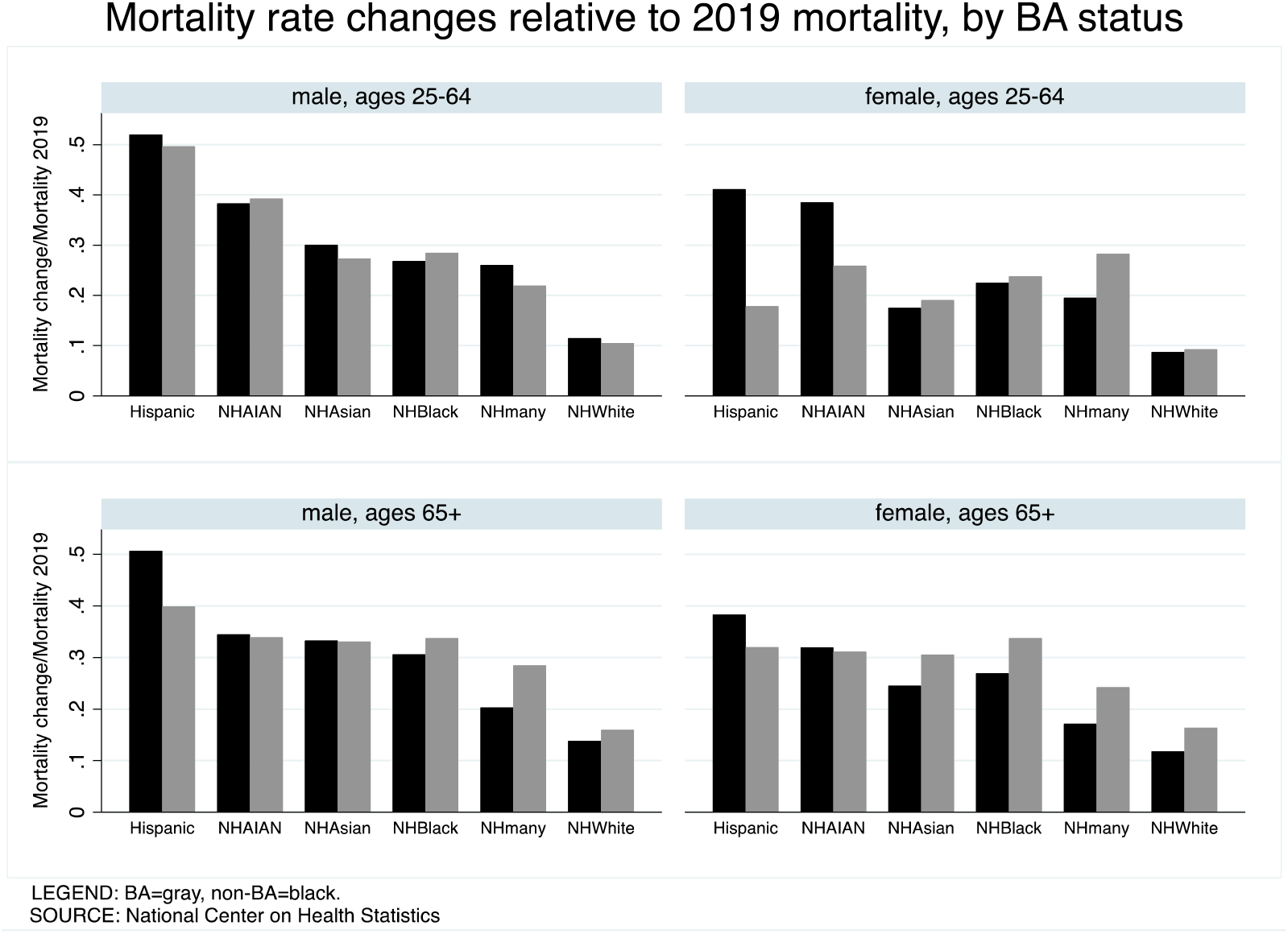

The most startling result in Figure 4 is the muted effects of having a BA on *relative* risk compared with the effects on *absolute* risk in Figure 3. For Hispanic and NHAIAN adult women, the relative risk is lower for those with a BA, as is true among Hispanic elderly men. But, for the large majority of groups, the relative risk was little different between those with and without a BA.

Figure 5 shows results for all five age groups in the underlying data, offering a check that the results are not contaminated by over-aggregation by age. The scatterplot shows the percentage protection associated with a BA in 2020 on the vertical axis versus the percentage protection associated with a BA in 2019. The solid line is the 45-degree line. In both years, the protective effect is larger for younger age groups. Beyond that, we see once again that, with a few exceptions discussed in the context of Figure 4, the efficacies of the BA were very little changed by the pandemic that, in other respects, did not treat people equally. There are 60 points in the figure; 32 are *negative*—a lower protective effect in 2020. The largest increases in protection are 10.0 percentage points (pp) for female NHAIAN aged 25-39, 9.5 pp for female Hispanics aged 55-64, and 9.5 pp for female Hispanics aged 25-39. The mean change is 0.23, confidence interval (−0.7, 1.1).

**Figure 5.**
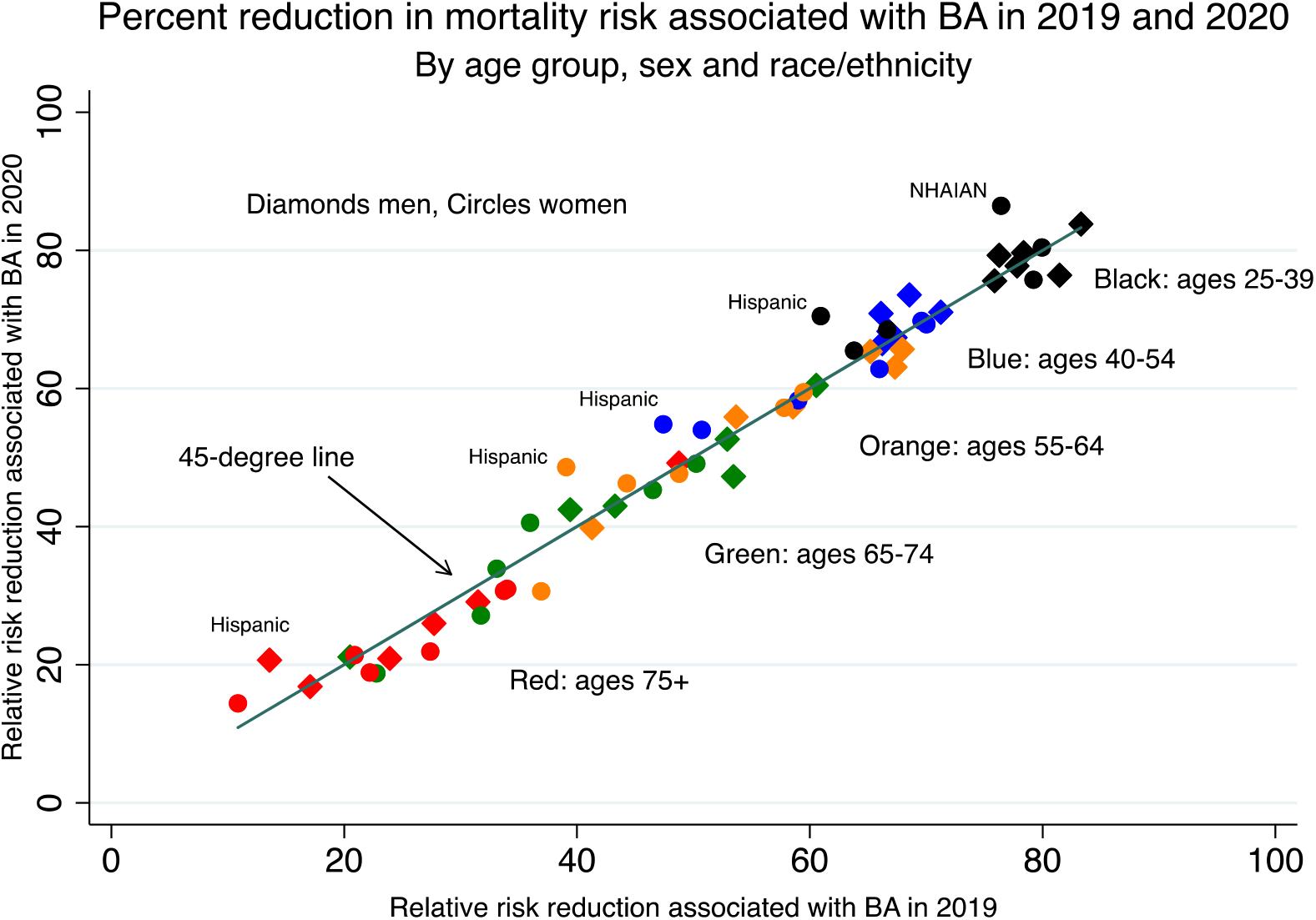

## 4. Discussion and conclusions

It is easy to understand why the BA degree was protective against COVID-19, and why the effect should be smaller for those aged 65 and above who are largely not in the labor force. People with and without the degree work in different occupations with different risks of infection. Those with a BA were more than twice as likely to be able to telework (65 versus 30 percent) and were twice as likely to be in a low-contact occupation (40 versus 20 percent) compared to those without, Daly et al (2020). Few frontline workers have a BA, and frontline workers were more likely to die, Chen YH et al (2021). Sixteen percent of workers in grocery, convenience, and drug stores had a BA, 15 percent in public transit, 10 percent in trucking, warehouse, and postal service, and 9 percent in building cleaning services, occupations that are also structured by race/ethnicity and gender, Rho et al (2020).

An exception to frontline workers being disproportionately drawn from the ranks of the less educated is health care, where more than 40 percent of workers have a BA or more. Healthcare workers were three times more likely to die than all workers. Even so, in the first year of the pandemic, more than two-thirds of healthcare workers who died were people of color, and “(l)ow paid workers who handled everyday patient care, including nurses, support staff and nursing home employees, were far more likely to die in the pandemic than physicians were,” Kaiser Health Network (2021).

These COVID risks for frontline workers were new in 2020, and were not simply increases in pre-existing risks, let alone proportional increases.

Irrespective of occupation, less-educated Americans have lower incomes, which increases the risk of infection, for example through crowded lower-quality housing making social distancing more difficult. Intrahousehold infection was important, Lee et al (2020). Our own calculations from the American Community Survey show that NHW and NHB household sizes do not vary by BA status, but for Hispanics and other NH groups, adults with a BA live in smaller households on average. Long-standing patterns of racial and ethnic segregation affect who lives where, the quality of local medical care, Bach et al (2004), and who rides on public transit, which likely caused many infections, particularly early in the pandemic in New York, Harris (2020), Kissler et al (2020), Sy et al (2021). Education was also related to compliance with public health recommendations in 2020, Weiss and Paasche-Orlow (2020).

The real mystery is not why the BA was so protective during the pandemic, but why the effect was almost as large *before* the pandemic. Or put differently, all of the arguments above point to an *increase* in the protective effect of a BA during the pandemic, that the gray bars in Figure 4 should be lower than the black bars, and that all the points in Figure 5 should be above the 45-degree line, neither of which happened. The occupations for less-educated Americans that were so dangerous during the pandemic were not as dangerous before the pandemic, so that the protection provided by the BA should have increased, and, with few exceptions, it did not.

In spite of a large literature, we do not fully understand the links between education and health, nor why the protective effect of a BA has been rising in the US in recent years. That education is associated with better health has been documented in many countries at many times. There are plausible mechanisms with causality running from education to health behaviors and health, from health—especially childhood health—to education, and mechanisms in which third factors influence both, Cutler and Lleras-Muney (2006). The college wage premium—the percent by which wages of those with a four-year degree exceed those with a high school diploma—has doubled since 1980, from 40 percent to 80 percent, James (2012), paralleling the rise in the mortality premium in Figure 2. As documented elsewhere, Case and Deaton (2020), the epidemic of deaths of despair is both worse and getting worse among younger Americans, and is much less serious among those with a four-year degree. Case and Deaton (2020) argue that the fifty-year long dysfunction of the American labor market for less-educated Americans has undercut the pillars of a successful life, weakening or destroying the institutions of community, marriage, and religion on which working people depend, as well as reducing their wages and their attachment to employment. Our results here suggest that the COVID-19 pandemic in 2020, far from being a break with these previous trends, is a continuation of them in a new disease environment with the fundamental inequalities persisting.

A partial and speculative explanation comes from decomposing the change in relative risks for those with and without a BA into the sum of the change in relative risk of dying conditional on infection and the change in relative risk of infection. The discussion above focuses on the latter. But the former depends on comorbidities—such as obesity, diabetes, alcohol use disorder, or COPD— which, prior to the pandemic, were patterned by age, race/ethnicity and education, Pampel et al (2010).

More formally, for any given subpopulation, we can write the mortality rate *m* as the product of the probability of dying, conditional on infection, i.e. the infection fatality rate (IFR), multiplied by the probability of infection *p*. In logarithmic terms

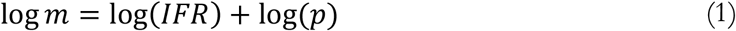

For two groups, 1 and 0, say those with and without the BA degree, we then have

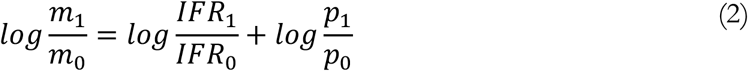

The discussion above supports the expectation that the second term on the right-hand side of (2) should have fallen from 2019 to 2020, while our results show that, for most groups, the left-hand side of (2) did not change. It is possible that risk factors—age, comorbidities, or lack of education— generated the same relative risk prior to and after COVID, which would be the case if COVID simply scaled up pre-existing risks. Given that the relative mortality rates changed little between the two years, equation (2) implies that the second term on the right-hand side is small or is less important than expected.

We conclude by noting a number of weaknesses in this study. The data are provisional, and depend on correct identification of race/ethnicity on death certificates. Racial/ethnic classification at death may differ from self-reported classifications in the American Community Survey. This is particularly problematic for AIAN, Arias et al (2016). Education may be incorrectly recorded, although the BA/non-BA distinction on death certificates is likely more accurate than that for high-school completion, Rostron et al (2010). Our results compare 2020 with 2019, and may not apply in other settings. We are not estimating parameters, but documenting a puzzle using notionally (if provisional) complete counts of deaths, so that, apart from the population estimates that are denominators in Figure 3 (but not in Figures 4 or 5, which depends on death counts only), they are not subject to standard errors or, more precisely, have standard errors of zero in the appropriate finite population calculation.

## Data Availability

Publicly available data, with a link provided in the text.

https://data.cdc.gov/NCHS/AH-Deaths-by-Educational-Attainment-2019-2020/4ueh-89p9

https://usa.ipums.org/usa/

https://www.cdc.gov/nchs/data_access/vitalstatsonline.htm

## Author statement

Anne Case: Conceptualization, Methodology, Data curation, Writing- Original draft preparation. Visualization, Investigation. Angus Deaton: Conceptualization, Methodology, Data curation, Writing-Original draft preparation. Visualization, Investigation.

## Acknowledgements

The authors gratefully acknowledge funding from the National Institute on Aging, R01AG053396 and R01AG060104. The NIA had no role in study design; in the collection, analysis and interpretation of data; in the writing of this article; or in the decision to submit it for publication.

